# Exploratory Assessment of Pulsed-Wave Doppler Representations of Lung Sounds Using Deep Learning: An In-Vitro Phantom Study

**DOI:** 10.64898/2026.06.09.26353787

**Authors:** Ali A. Saad, Sarah B. Murthi, Emad M. Boctor, William A. Teeter, Nitin Seam

## Abstract

The increasing availability of portable ultrasound systems motivates exploration of novel approaches to respiratory signal assessment. In this in-vitro study, we investigate whether pulsed-wave (PW) Doppler ultrasound can capture structured spectral patterns from replayed lung sound recordings.

Digitized respiratory sounds were replayed through a tissue-mimicking ultrasound phantom, generating 1,478 PW Doppler spectral images from recordings associated with healthy subjects and several externally labeled disease categories. Exploratory classification experiments using a ResNet-18 architecture demonstrated that these Doppler representations contain learnable differences under controlled conditions.

These findings motivate further investigation into PW Doppler as a potential representation of respiratory acoustics.

## Introduction

Auscultation with a stethoscope remains the standard bedside tool for examining the respiratory system, providing a rapid and inexpensive method to assess airflow and detect abnormal breath sounds such as wheezes and crackles [1][2].

However, lung auscultation has important limitations, including modest diagnostic sensitivity and substantial interobserver variability, even among trained clinicians. Prior studies and meta-analyses have demonstrated inconsistent recognition and interpretation of respiratory sounds, limiting the reliability of auscultation for disease classification [3–6].

To address these limitations, digital stethoscopes and artificial intelligence–based approaches have been increasingly explored to enhance respiratory sound acquisition, standardization, and automated classification, with several studies demonstrating improved agreement with expert assessment and potential clinical utility [7–10].

On the other hand, lung ultrasound (LUS) has become a core bedside imaging modality across emergency, intensive care, and general medical settings, delivering real-time, radiation-free assessment and frequently complementing—or even surpassing—chest radiography for common pulmonary conditions, and is increasingly recognized as an important modality for imaging the lung and pleura in critical care [11][12]. Robust evidence and consensus statements support LUS for rapid diagnosis of pneumonia, pleural effusion, pneumothorax, and pulmonary edema [13][14]. Protocolized approaches, such as the BLUE protocol, have standardized the evaluation of acute respiratory failure and improved diagnostic accuracy in critical care [15]. LUS is also used for procedural guidance during thoracentesis and other pleural interventions [12][15]. Recent international guidelines emphasize that LUS achieves high sensitivity with comparable specificity to chest X-ray for pneumonia and other conditions [13][14], is repeatable at the bedside, and integrates seamlessly into point-of-care workflows—including handheld devices and tele-ultrasound [16]. The versatility and safety profile of LUS have led to its rapid adoption in both adult and pediatric populations, and ongoing advances continue to expand its clinical utility [17][18].

Recent research has explored the diagnostic overlap between traditional pulmonary auscultation using the stethoscope and lung ultrasound imaging. While auscultation remains a cornerstone of bedside respiratory assessment, its limitations—such as poor sensitivity, interobserver variability, and technical challenges in noisy or critical care environments—are well known [19]. Lung ultrasound has been found to be superior to auscultation in diagnosing pulmonary edema and critically ill patients and other indications [20].

To our knowledge, prior work has not systematically evaluated whether pulsed-wave Doppler ultrasound can provide a meaningful representation of respiratory sound signals. Accordingly, the goal of this study was not to establish a diagnostic method, but rather to address this question using a controlled in-vitro phantom setup, in which PW Doppler spectral patterns are generated from replayed digitized lung sound recordings and examined for learnable structure. If such a structure exists, it may motivate future investigation into its underlying physiologic basis and whether similar representations can be observed in vivo.

## Methodology

We describe the methodology of an in-vitro study designed to explore whether pulsed-wave Doppler ultrasound can generate distinct spectral representations from replayed respiratory sound recordings associated with different annotated sound and disease categories.

### Lung Sounds Database

The lung sounds database [21] utilized in this study was developed for the ICBHI 2017 Scientific Challenge and consists of real-world lung auscultation recordings collected over several years from patients in hospitals in Portugal and Greece, using electronic and acoustic stethoscopes, with respiratory cycles annotated by clinical experts for wheezes, crackles, both, or normal sounds. The database also includes subject-level disease labels that were derived from associated clinical records or study metadata (e.g., spirometry for COPD, or imaging for pneumonia). These labels were not generated using a standardized adjudication protocol across all collection sites and should therefore be considered heterogeneous clinical descriptors rather than gold-standard diagnostic ground truth.

The database consists of 5.5 hours of audio recordings from 126 patients, including 6,898 respiratory cycles annotated for crackles and wheezes. It contains 920 audio files, each labeled by respiratory experts for adventitious sounds, representing a spectrum of respiratory conditions, including healthy controls, chronic obstructive pulmonary disease (COPD), asthma, upper respiratory tract infection (URTI), lower respiratory tract infection (LRTI), bronchiectasis, and pneumonia. In the present study, these disease categories were used only as coarse external groupings for exploratory analysis. Each patient contributed multiple recordings, with lung sounds captured from various anatomical chest locations—specifically the trachea (Tc), as well as anterior (A), posterior (P), and lateral (L) regions on both the left (l) and right (r) sides. This comprehensive sampling ensures coverage of clinically relevant auscultation sites for respiratory sound analysis.

For each audio file, detailed metadata were recorded, including the chest location, acquisition mode (sequential/single channel [sc] or simultaneous/multichannel [mc]), and the type of recording equipment used. Devices included the AKG C417L Microphone, 3M Littmann Classic II SE Stethoscope, 3M Littmann 3200 Electronic Stethoscope, and Welch Allyn Meditron Master Elite Electronic Stethoscope. Importantly, all recordings for a given patient were acquired using the same equipment and acquisition mode to maintain consistency across chest locations. This rich dataset supports exploratory investigation of signal representations for automated lung sound analysis in controlled experimental settings. Accordingly, the database was used in this study to support in-vitro exploration of pulsed-wave Doppler representations of replayed lung sounds, rather than to establish diagnostic performance.

### Phantom Construction

To use the lung sounds database with Doppler ultrasound imaging, we constructed a phantom for this purpose, as shown in Figure 1 below. A HEYSONG Waterproof Bluetooth Speaker (Model B0B3DN72R8) [22] was used as the acoustic source in the experimental setup. The speaker was connected via Bluetooth to a computer and used to reproduce digital lung sound files from the selected database, ensuring accurate playback of adventitious lung sounds. The phantom consisted of a Ylucky Ultrasound IV Training Phantom (Model B0CK3PNB3T) [23], constructed from a soft, tissue-mimicking material in a rectangular block form, designed to approximate key acoustic properties of soft tissue for realistic ultrasound and Doppler measurements. This phantom setup was intended to provide a controlled and repeatable environment for replaying respiratory sound recordings and characterizing their corresponding pulsed-wave Doppler responses, rather than to reproduce in-vivo thoracic biomechanics.

**Figure 1.**
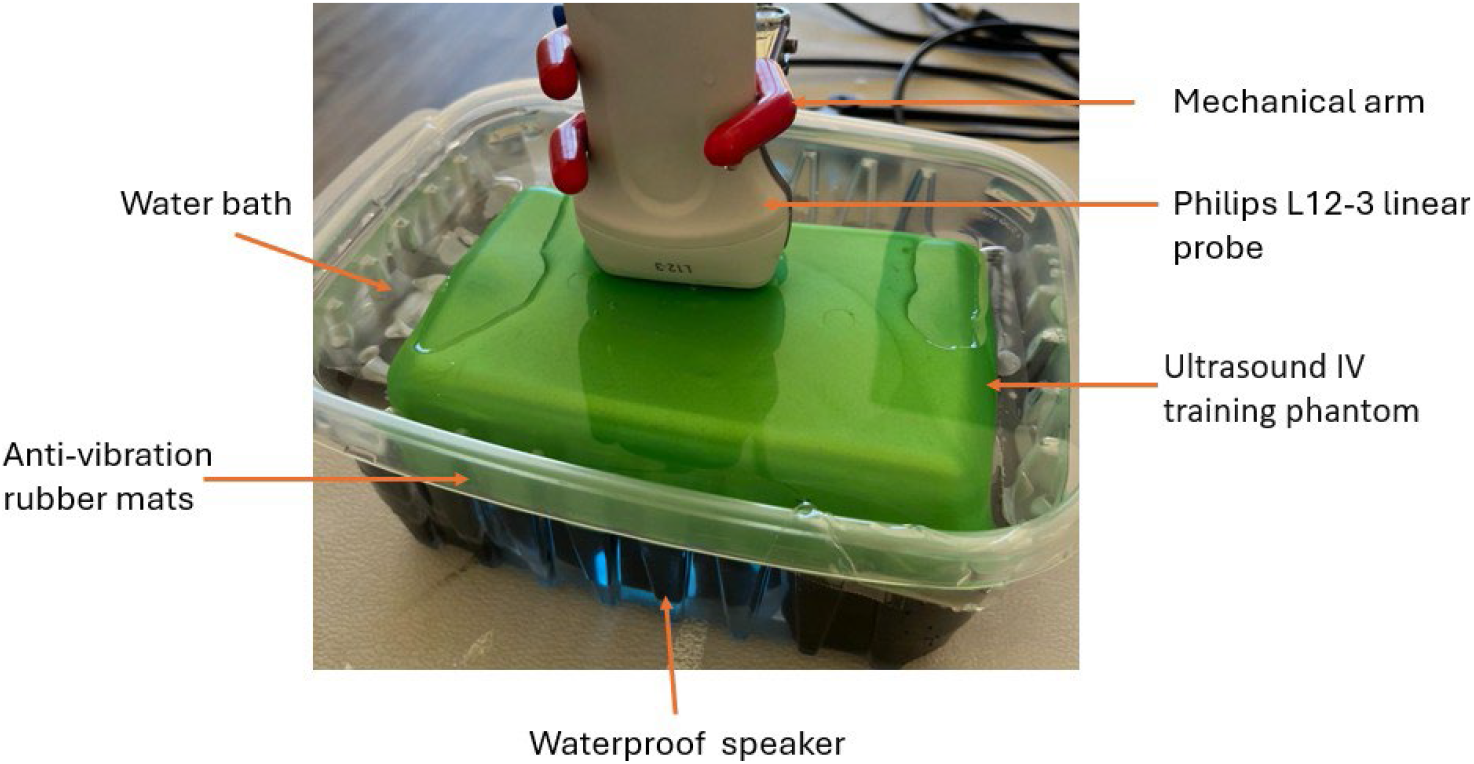
Lung sounds imaging phantom construction.

For the experiment, the phantom was placed above the waterproof speaker, with a water gap in between, and both components were fully submerged in a plastic water bucket to provide optimal acoustic coupling and minimize air interference. XCELERATOR Anti-Vibration Pads (Model B07FKVH8SN) [24], made of durable rubber, were positioned on the sides of the bucket to support and stabilize both the phantom and the speaker, effectively reducing external vibrations and ensuring measurement consistency.

A Philips Cx50 ultrasound system (Philips Medical Systems, Bothell, WA, USA) [25], shown in Figure 2, equipped with an L12-3 linear array transducer [26] was used to acquire pulsed wave (PW) Doppler images of the transmitted lung sound signals. The probe was mounted above the phantom and coupled through the water bath, allowing for Doppler characterization of motion-related signal components induced by the replayed acoustic waves within the tissue-mimicking phantom. All Doppler measurements therefore represent responses to replayed acoustic stimuli under controlled experimental conditions, rather than native physiologic signals generated by the respiratory system.

**Figure 2.**
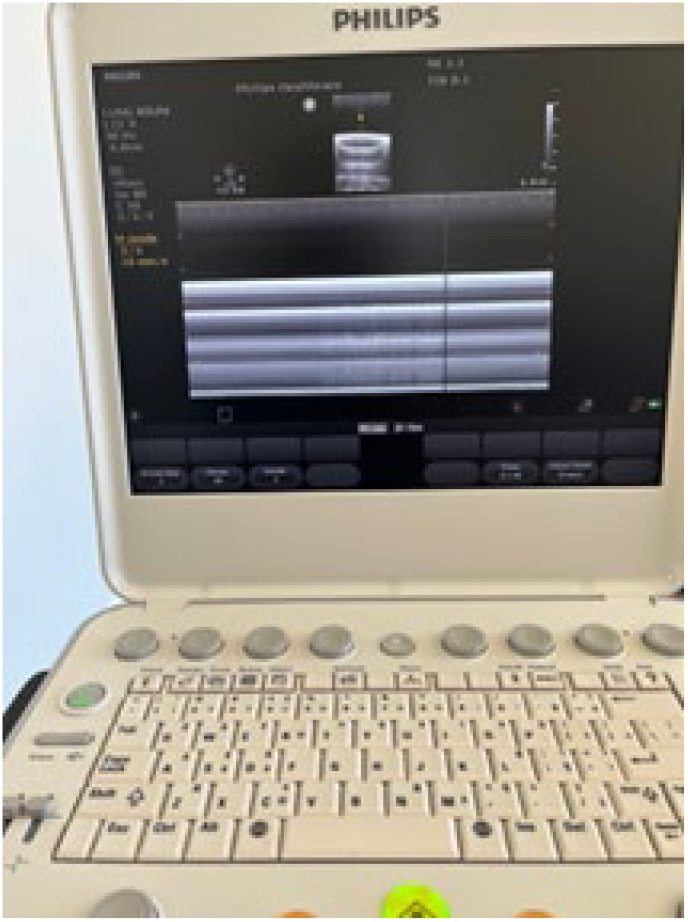
Philips Cx50 ultrasound system used for Doppler image captures.

### PW Doppler Image Collection & Preprocessing

The Philips Cx50 ultrasound system was configured to capture pulsed-wave (PW) Doppler spectrums generated in response to replayed lung sound files. A dedicated clinical preset was developed for these experiments. The Doppler angle was set to 0° to align with the vertical vibration direction of the speaker, ensuring optimal sensitivity for Doppler signal detection within the controlled phantom environment. The PW Doppler scrolling rates used were 10mm/s and 25mm/s. The Doppler scale was configured to optimize spectral signal representation and avoid aliasing, while the sample volume was positioned at the center of the tissue-mimickingphantom.

Each lung sound file from the database was used to create a corresponding study, named according to the original file, which encodes patient ID, chest location, acquisition mode, and recording equipment type. Most audio files were approximately 23 seconds in duration and fit within a single PW Doppler image. For longer recordings exceeding this duration, multiple non-overlapping PW Doppler images were acquired. The speaker volume was tested at 100% and 50% for each audio input to assess signal robustness under differing playback amplitudes. Figure 3 illustrates an example of PW Doppler acquisition from a replayed lung sound recording labeled as COPD. In total, 1478 PW Doppler images were collected from recordings associated with 126 subjects across various disease categories, as summarized in Table 1.

**Table 1.** Summary of PW Doppler images generated from replayed lung sound recordings by externally labeled categories.

| Disease | Number of Patients | Number of PW Doppler Images |
| --- | --- | --- |
| Healthy | 26 | 322 |
| COPD | 64 | 322 |
| Pneumonia | 6 | 335 |
| Bronchiectasis | 7 | 142 |
| Bronchiolitis | 6 | 138 |
| URTI | 14 | 181 |
| LRTI | 2 | 26 |
| Asthma | 1 | 12 |
| Total | 126 | 1478 |

**Figure 3.**
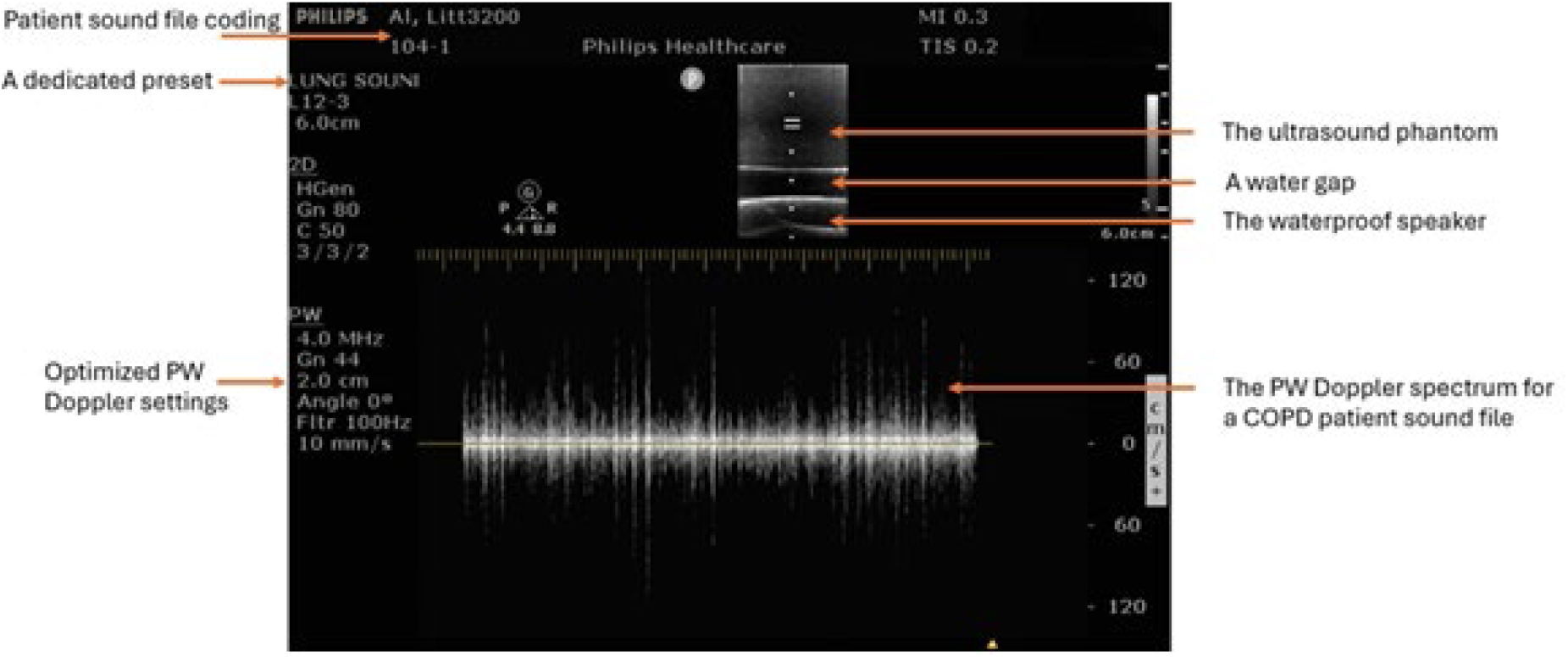
PW Doppler data acquisition example for a replayed lung sound recording labeled as COPD.

All images were exported in BMP format and transferred to a computer for machine learning analysis. Preprocessing involved custom MATLAB scripts to isolate the PW Doppler region from the spectral display for subsequent feature extraction. Figure 4, Figure 5, and Figure 6 present representative PW Doppler spectrums for replayed lung sound recordings labeled as healthy, COPD, and URTI, respectively. All PW Doppler images therefore represent responses to replayed acoustic stimuli acquired under controlled experimental conditions, rather than in-vivo physiologic measurements.

**Figure 4.**
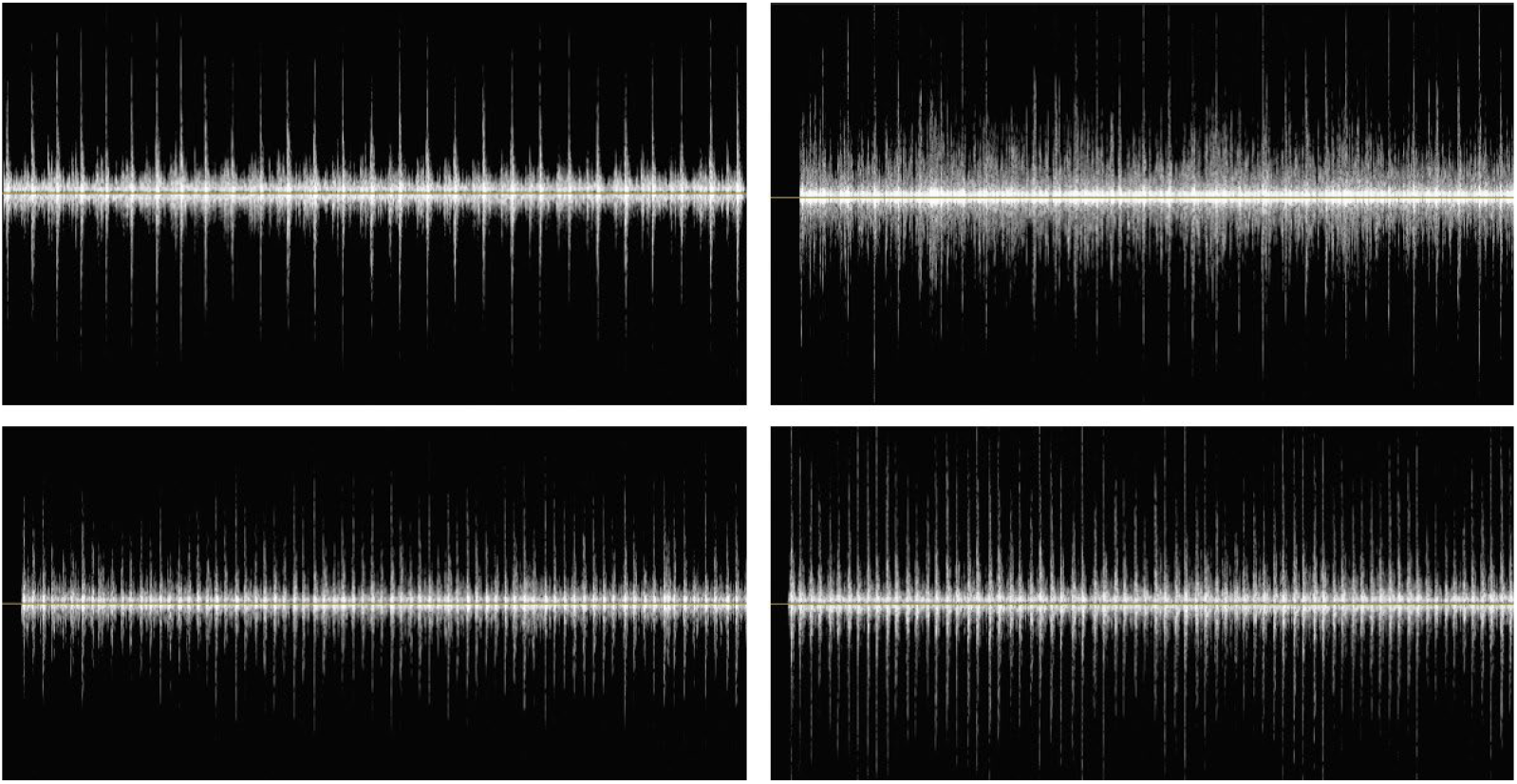
Representative PW Doppler acquisition examples from replayed lung sound recordings labeled as healthy.

**Figure 5.**
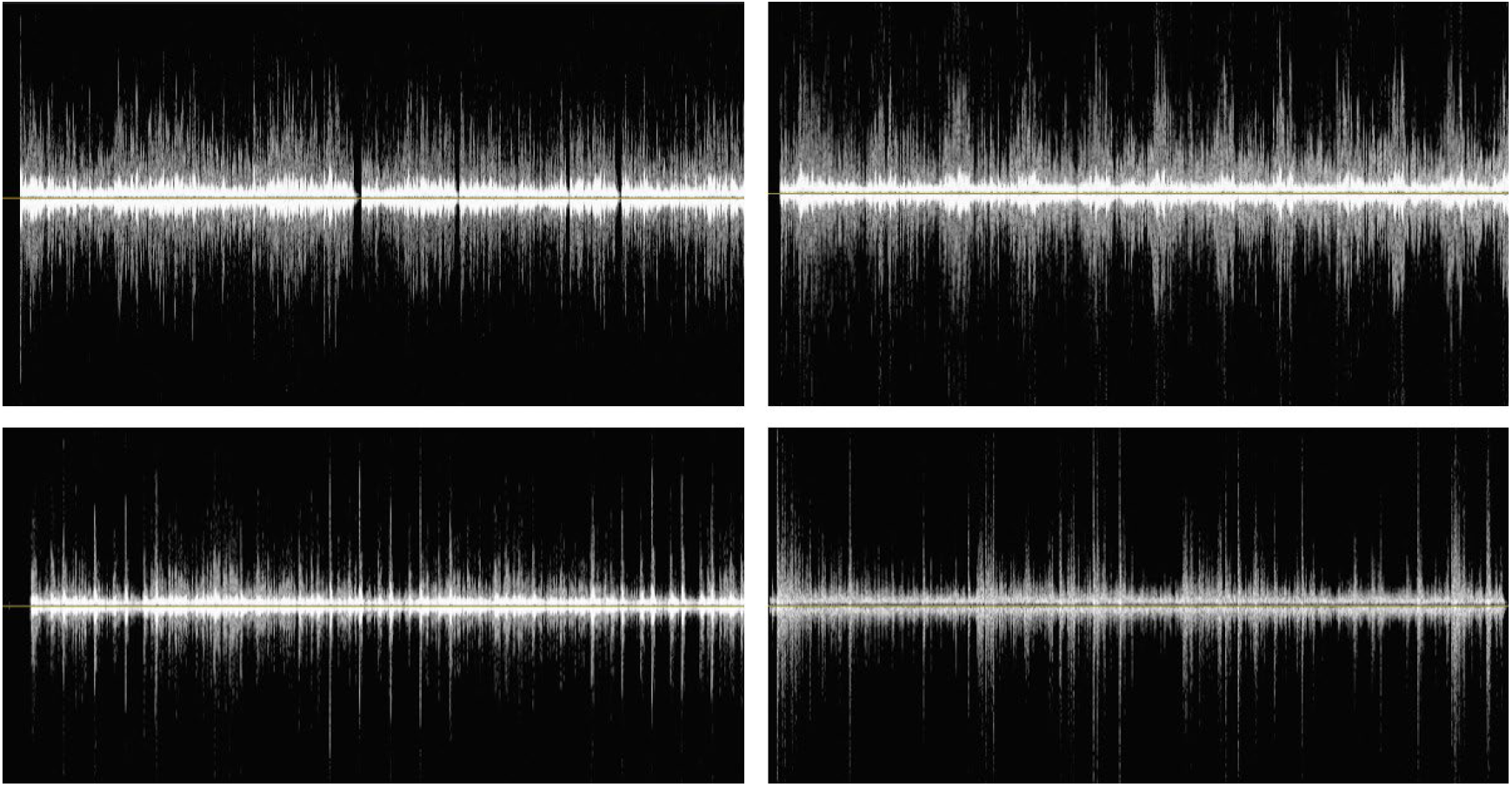
Representative PW Doppler acquisition examples from replayed lung sound recordings labeled as COPD.

**Figure 6.**
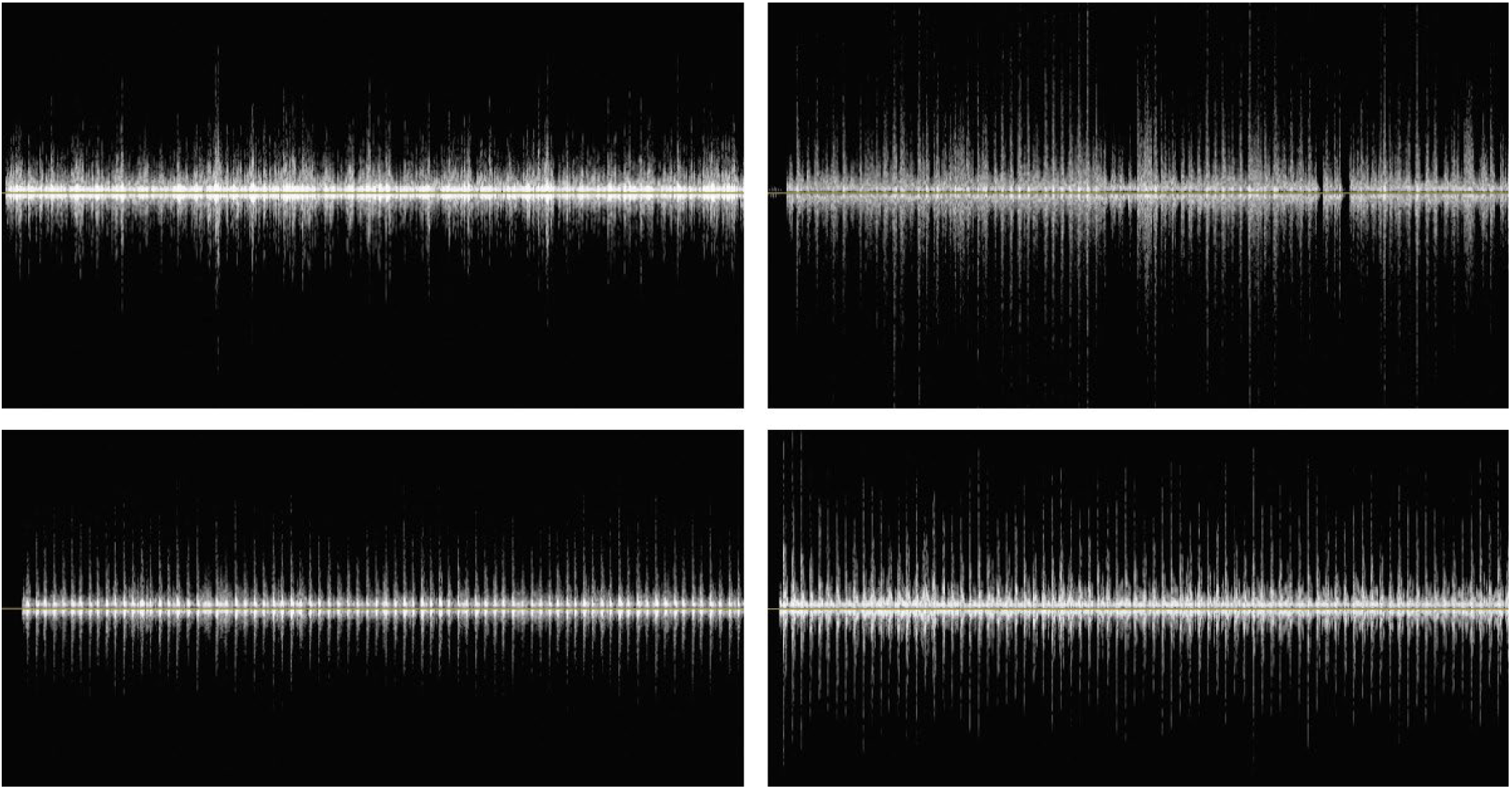
Representative PW Doppler acquisition examples from replayed lung sound recordings labeled as upper respiratory tract infection (URTI).

### Machine Learning Implementation

To support exploratory analysis of PW Doppler spectral images, we utilized MATLAB’s Deep Network Designer [27] as the primary platform for designing and configuring deep neural network architectures. The application provides an interactive graphical interface that enables researchers to construct network architectures layer by layer, visualize model structures, and set training parameters without requiring extensive programming. To optimize performance and reduce computational overhead, the methodology incorporated pretrained convolutional neural networks (CNNs) available within MATLAB’s deep learning toolbox [28].

The pretrained models utilized in this research were trained on the ImageNet dataset [29][30], a comprehensive visual database containing millions of labeled images across thousands of object categories.

The ResNet-18 model, introduced by He et al. (2016) [31], was selected for this research due to its balance between computational efficiency and accuracy.

Three exploratory image-level grouping experiments were conducted using a ResNet-18 network in MATLAB on the PW Doppler spectral images. These experiments included binary grouping analyses (Healthy-labeled vs. COPD-labeled recordings; Healthy-labeled vs. pooled disease-labeled recordings) as well as a multi-class grouping analysis. In each experiment, 50% of the images were used for training, 10% for validation, and 40% for testing, with partitioning performed at the image level. Figure 7 shows the network training settings used for all experiments.

**Figure 7.**
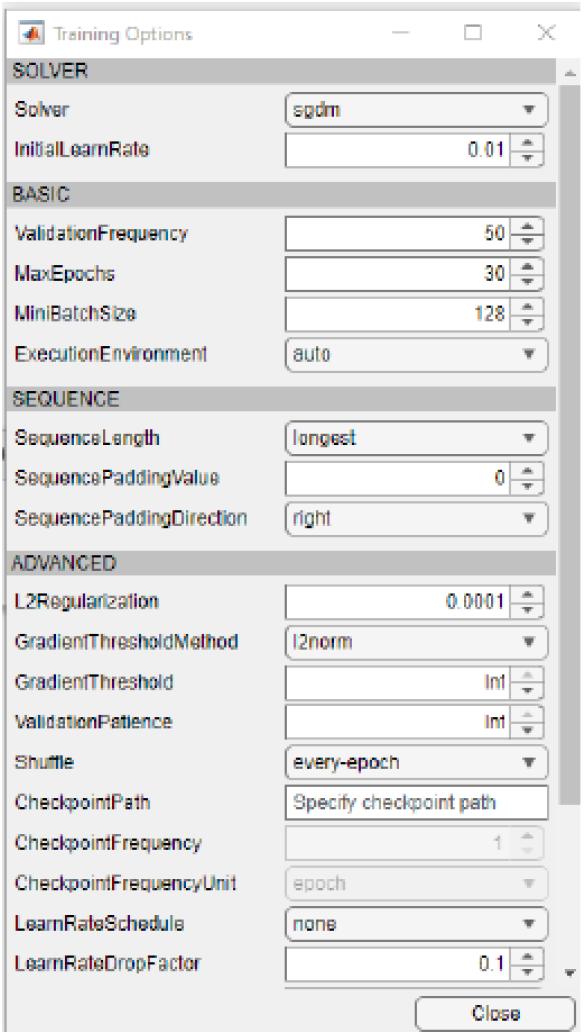
ResNet-18 training settings used for exploratory experiments.

The first experiment aimed to separate PW Doppler images derived from recordings labeled as COPD from those labeled as Healthy. The experiment was conducted using a balanced dataset comprising 322 COPD-labeled images and 322 Healthy-labeled images.

The second experiment aimed to separate Healthy-labeled images from images aggregated across externally labeled disease categories. This experiment was conducted using a balanced dataset comprising 322 images aggregated from all disease categories and 322 Healthy-labeled images.

In addition to the binary experiments, a multi-class grouping analysis was conducted to assess separability among multiple externally labeled category groups. The experiment included five disease categories—COPD, bronchiectasis, bronchiolitis, pneumonia, and upper respiratory tract infection (URTI)—along with the Healthy class. To mitigate class imbalance and enable consistent comparison across groups, a balanced dataset was constructed by selecting an equal number of PW Doppler images (138 images) from each category, including Healthy.

For the multi-class classification, performance was assessed using a one-vs-rest evaluation strategy for each disease class, with the Healthy-labeled class treated exclusively as a negative reference when reporting disease-labeled specific metrics at the image level.

Performance metrics and confusion matrices were derived from the true positive (TP), true negative (TN), false positive (FP), and false negative (FN) values computed on the held-out image set. In this framework, the confusion matrix summarizes predicted versus true reference labels, with entries corresponding to TP, TN, FP, and FN, analogous to a standard diagnostic test table. These values allow direct calculation of clinically familiar performance measures, including sensitivity (true positive rate), specificity (true negative rate), and positive and negative predictive values (PPV and NPV), which reflect the likelihood that predicted classifications are correct. Reported metrics also included overall accuracy, balanced accuracy, false positive rate (FPR), and false negative rate (FNR). Figure 8 lists the performance metric calculations used.

**Figure 8.**
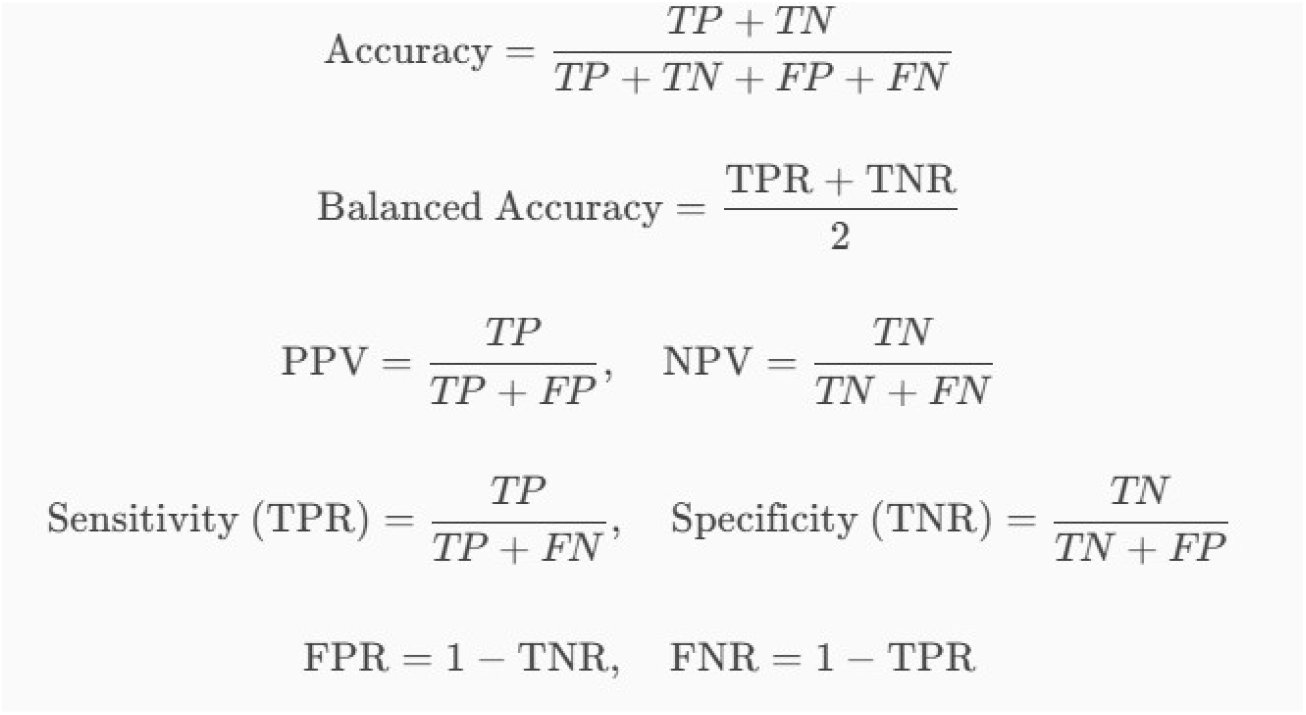
Classification metrics used for exploratory experiments.

## Results

For the first classification task (COPD-labeled images vs. Healthy-labeled images), the training plot is shown in Figure 9. The confusion matrix is shown in Figure 10, while the binary classification metrics are summarized in Table 2.

**Table 2.**
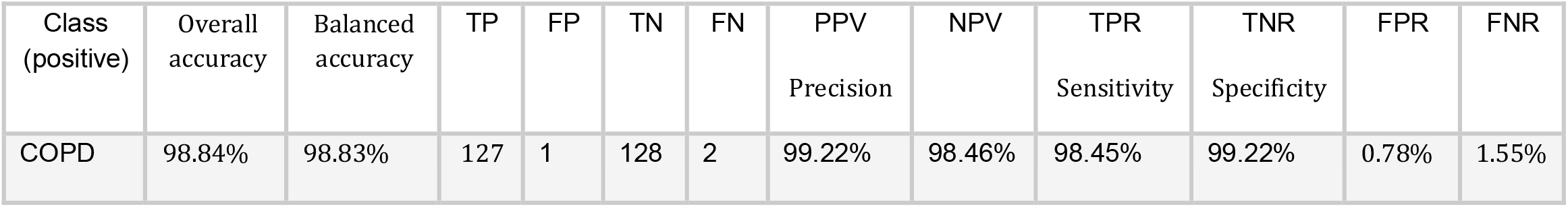
Performance metrics for the COPD-labeled versus Healthy-labeled image-level experiment.

**Figure 9.**
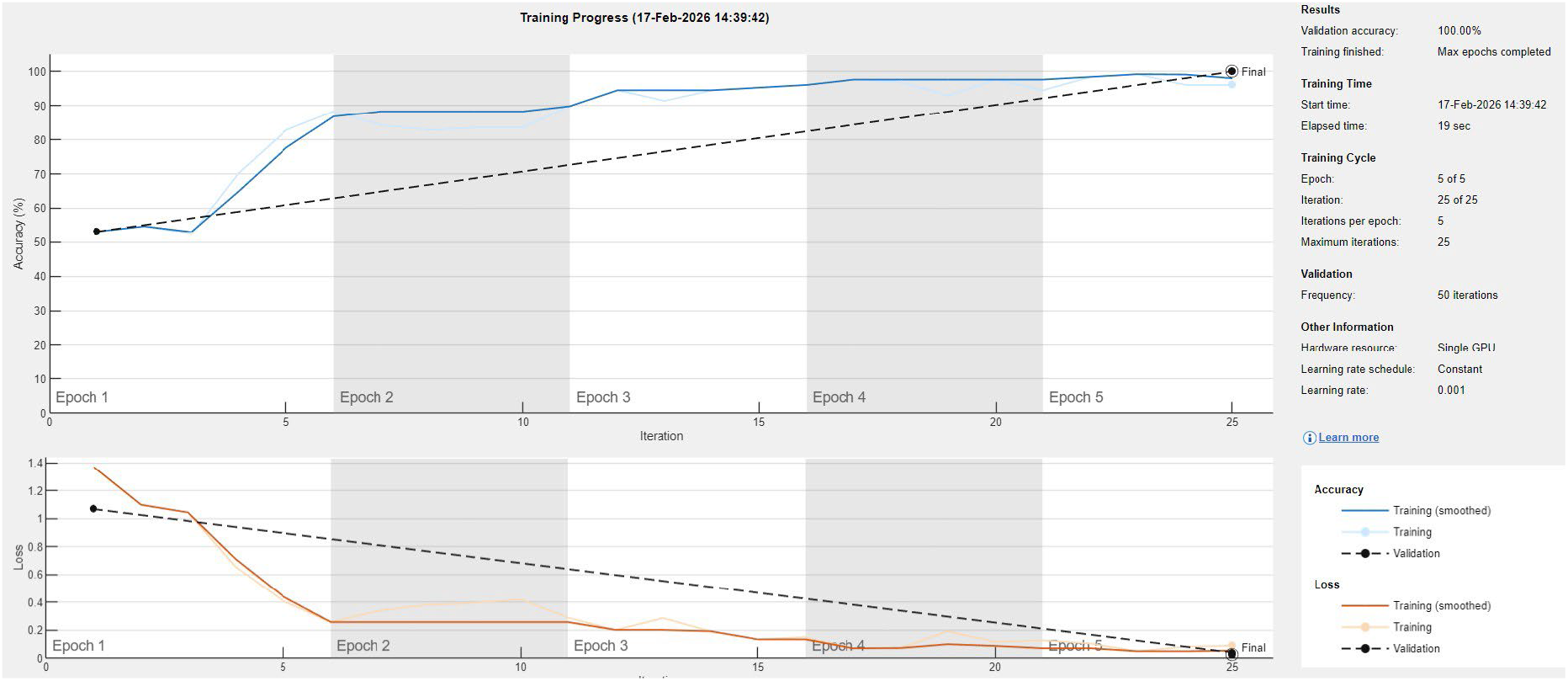
Training plot for the COPD-labeled vs. Healthy-labeled experiment.

**Figure 10.**
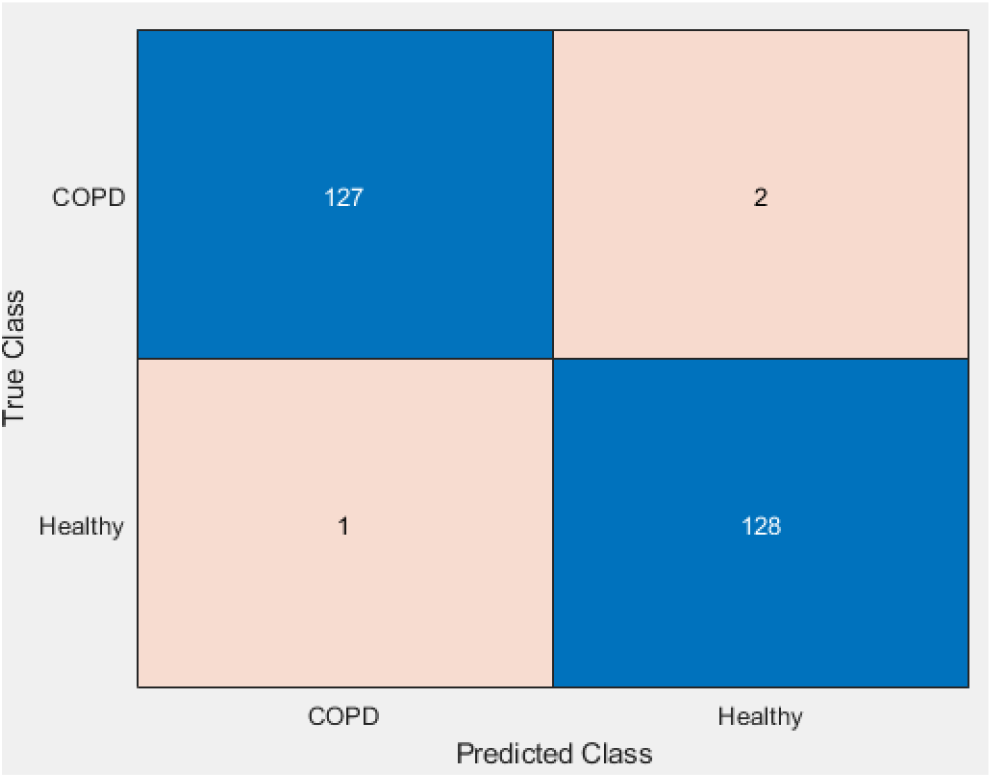
Confusion matrix for the COPD-labeled versus Healthy-labeled image-level experiment.

In the COPD-labeled versus Healthy-labeled image-level experiment, high separation was observed within the held-out test set. Of the 129 COPD-labeled images, 127 were correctly assigned, with two misclassified as Healthy-labeled images, while 128 of 129 Healthy-labeled images were correctly assigned. These results indicate that PW Doppler spectral images derived from replayed lung sound recordings contain differentiable patterns under controlled experimental conditions.

The confusion matrix for the second exploratory image-level grouping experiment (Healthy-labeled vs. pooled disease-labeled recordings) is shown in Figure 11, with corresponding metrics summarized in Table 3.

**Table 3.**
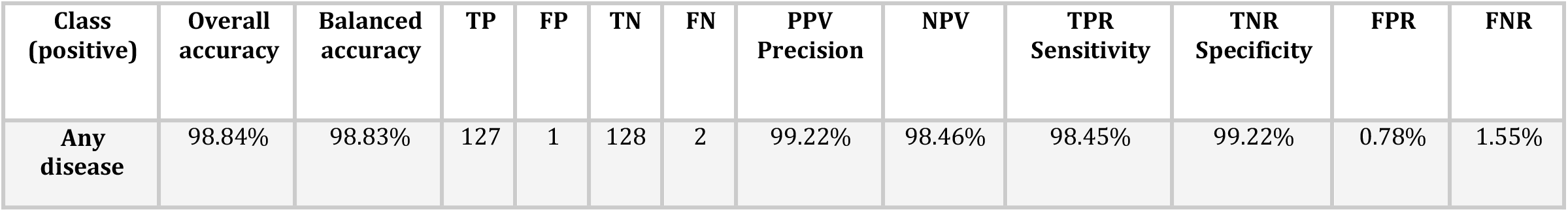
Performance metrics for the Healthy-labeled versus pooled disease-labeled image-level experiment.

| Class (positive) | Overall accuracy | Balanced accuracy | TP | FP | TN | FN | PPV Precision | NPV | TPR Sensitivity | TNR Specificity | FPR | FNR |
| --- | --- | --- | --- | --- | --- | --- | --- | --- | --- | --- | --- | --- |
| Any disease | 98.84% | 98.83% | 127 | 1 | 128 | 2 | 99.22% | 98.46% | 98.45% | 99.22% | 0.78% | 1.55% |

**Figure 11.**
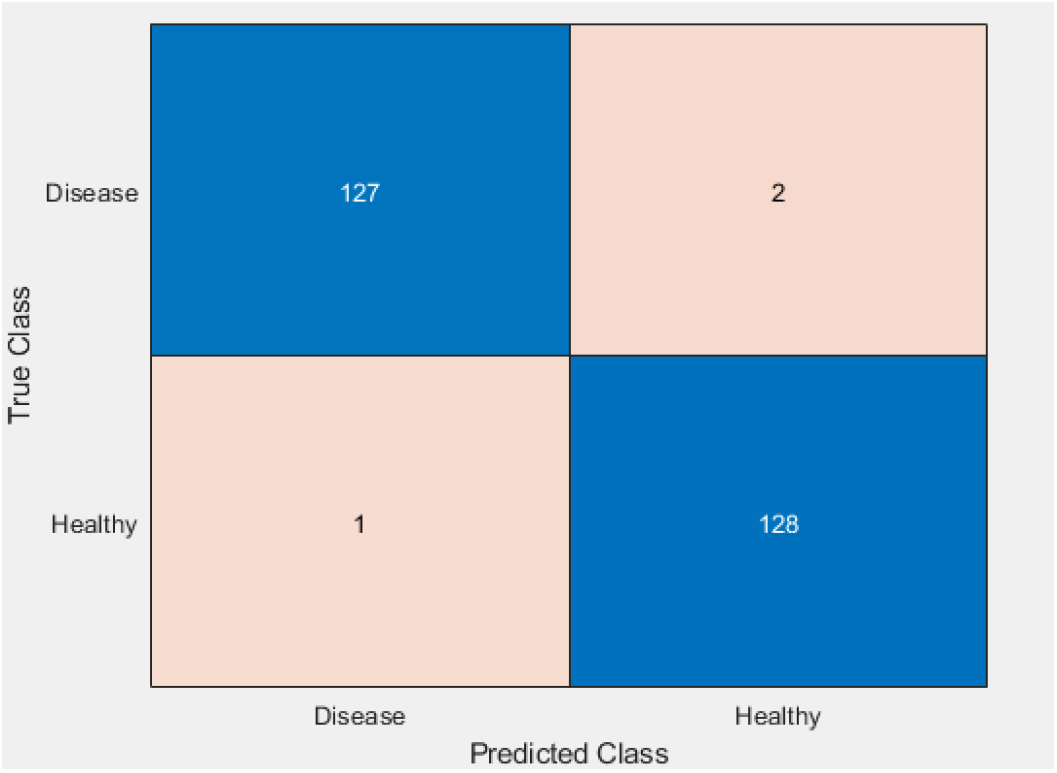
Confusion matrix for the Healthy-labeled versus pooled disease-labeled image-level experiment.

In the Healthy-labeled versus pooled disease-labeled image-level experiment, strong separation was again observed within the held-out test set. Of the pooled disease-labeled images, 127 of 129 were correctly assigned, while 128 of 129 Healthy-labeled images were correctly assigned. These results suggest that aggregated disease-associated recordings produce PW Doppler spectral representations that differ from those associated with healthy recordings in this controlled in-vitro setting.

The confusion matrix for the third classification task (multi-classification task) is shown in Figure 12, and the standard classification metrics (shown in Table 4), including sensitivity, specificity, precision, and accuracy, were derived to characterize per-class performance for exploratory analysis.

**Table 4.** Performance metrics for the multi-class image-level grouping experiment.

| Class | TP | FP | TN | FN | PPV<br>Precision | NPV | TPR<br>Sensitivity | TNR<br>Specificity | FPR | FNR | Accuracy |
| --- | --- | --- | --- | --- | --- | --- | --- | --- | --- | --- | --- |
| **Bronchiectasis** | 52 | 3 | 272 | 3 | 94.55% | 98.91% | 94.55% | 98.91% | 1.09% | 5.45% | 98.18% |
| **Bronchiolitis** | 51 | 2 | 273 | 4 | 96.23% | 98.56% | 92.73% | 99.27% | 0.73% | 7.27% | 98.18% |
| **COPD** | 54 | 1 | 274 | 1 | 98.18% | 99.64% | 98.18% | 99.64% | 0.36% | 1.82% | 99.39% |
| **Pneumonia** | 55 | 2 | 273 | 0 | 96.49% | 100.0% | 100.0% | 99.27% | 0.73% | 0.0% | 99.39% |
| **URTI** | 53 | 3 | 272 | 2 | 94.64% | 99.27% | 96.36% | 98.91% | 1.09% | 3.64% | 98.48% |

**Figure 12.**
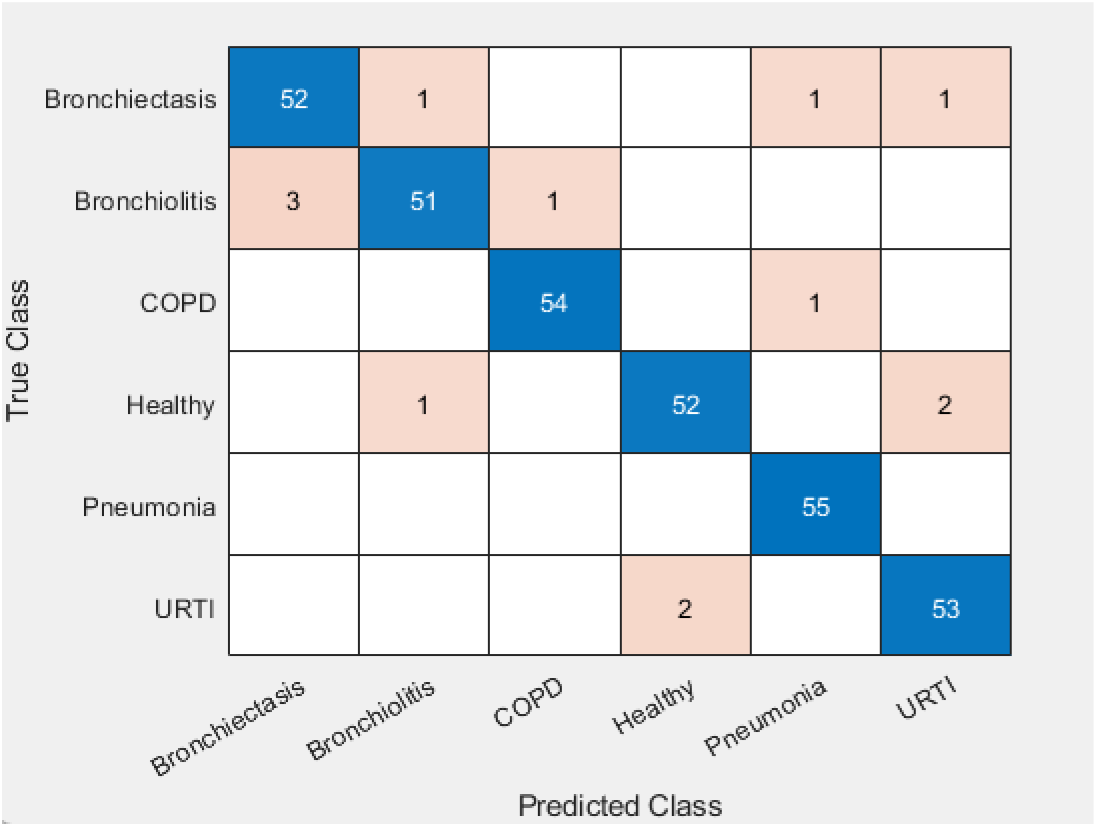
Confusion matrix for the multi-class image-level grouping experiment.

Using pulsed-wave Doppler ultrasound spectrum images, the proposed deep learning framework demonstrated separability across externally labeled category groups in a multi-class classification setting. Performance was assessed using a one-vs-rest approach for each disease category, with the Healthy-labeled image class treated exclusively as a negative reference and excluded as a positive class.

Across all externally labeled categories—Bronchiectasis, Bronchiolitis, COPD, Pneumonia, and URTI—the model achieved high image-level separation within the balanced test set. These results indicate that PW Doppler spectral representations derived from replayed lung sound recordings retain structured differences across the selected label groups.

## Discussion

This study explores the feasibility of using pulsed-wave Doppler ultrasound combined with deep learning to assess whether pulsed-wave Doppler spectral representations derived from replayed lung sound recordings are computationally differentiable in an in-vitro setting. The results from both binary and multi-class classification experiments indicate that Doppler spectral representations of lung sounds contain structured differences that can be learned effectively using convolutional neural networks.

Across the binary image-level grouping experiments (Healthy-labeled vs. COPD-labeled and Healthy-labeled vs. pooled disease-labeled recordings), strong separation was observed within the controlled in-vitro dataset. These results suggest that PW Doppler spectral representations of replayed lung sounds contain consistent patterns that can be computationally distinguished under experimental conditions. Importantly, these findings should not be interpreted as evidence that PW Doppler provides a diagnostic alternative to conventional auscultation, but rather that it may offer a complementary signal representation worthy of further investigation.

The multi-class experiment further demonstrates the potential of this approach to separate multiple externally labeled category groups at the image level, including COPD, bronchiectasis, bronchiolitis, pneumonia, and URTI. The multi-class experiment further illustrates that PW Doppler spectral representations encode structured variation across the included externally labeled categories. Given the limited number of subjects in several classes and the heterogeneous nature of the labels, these results are best interpreted as exploratory pattern separation rather than evidence of reliable disease-specific discrimination.

This work highlights a novel intersection between functional acoustic assessment and imaging-based diagnostics. While lung ultrasound is widely used for detecting structural abnormalities, its application to respiratory sound analysis has been limited. By transforming acoustic vibrations into visual Doppler spectrograms suitable for machine learning, this study introduces a hybrid paradigm that may complement existing lung ultrasound protocols from a research perspective.

Several limitations should be acknowledged. The experiments were conducted using an in-vitro phantom and replayed acoustic stimuli, which enables controlled and repeatable measurements but does not fully capture in-vivo anatomical complexity, respiratory motion, or physiological variability. In addition, classification was performed at the image level rather than patient-level inference, and future work will be needed to assess patient-level performance and clinical integration. As a feasibility study, these findings should be interpreted as preliminary, motivating further investigation rather than definitive clinical validation.

## Conclusion

In this in-vitro study, we explored the feasibility of using pulsed-wave Doppler ultrasound combined with deep learning to evaluate PW Doppler spectral representations of replayed adventitious lung sound recordings. Using a controlled phantom setup and a curated respiratory sound database, the proposed framework demonstrated consistent image-level separation across several exploratory grouping experiments.

These findings suggest that Doppler ultrasound–derived spectral representations of lung sounds contain discriminative information that can be effectively leveraged by convolutional neural networks. By transforming acoustic phenomena into visual Doppler patterns suitable for automated analysis, this approach offers a potential representation framework for studying respiratory acoustic signals in conjunction with ultrasound imaging.

Although these findings are based on in-vitro studies, they demonstrate that Doppler ultrasound warrants further exploration as a functional acoustic imaging method. This work establishes a technical foundation for future studies aimed at translating Doppler-based lung sound analysis into future physiologic and in-vivo investigations.

### Next Steps

Building on the feasibility demonstrated in this study, several directions for future work are warranted.

First, in-vivo validation studies are needed to evaluate the robustness of the proposed approach under realistic clinical conditions, including respiratory motion, chest wall variability, heterogeneous tissue properties, and ambient noise. Such studies will be essential for assessing generalizability beyond the controlled phantom environment.

Second, future investigations should focus on patient-level analysis, including aggregation of multiple Doppler images per subject and evaluation of longitudinal consistency. This will better align model outputs with clinical decision-making workflows.

Third, expanding the dataset to include a wider range of patients, varying disease severities, and different respiratory conditions—particularly those less represented such as asthma and LRTI—will improve multi-class performance and reduce bias.

Additional avenues include exploring temporal and sequence-based learning approaches to capture dynamic Doppler patterns over time, as well as evaluating deployment on portable and handheld ultrasound platforms for point-of-care and telemedicine applications.

Collectively, these next steps aim to transition Doppler ultrasound–based lung sound analysis from a controlled experimental setting toward real-world clinical feasibility evaluation.

## Data Availability

The public lung sounds database used in this study can be found here: https://ai4eu.dei.uc.pt/respiratory-sounds-dataset/
All other data produced in the present study are available upon reasonable request to the authors.

## Acknowledgements

The authors used an AI-assisted tool to support literature exploration, language refinement, and manuscript organization. All sources were independently reviewed by the authors, and all scientific content, analysis, and conclusions were verified and approved by the authors, who take full responsibility for the work.

